# Screen-based media device use, its associated factors and sleep outcomes among undergraduate health students studying in colleges of Kathmandu valley, Nepal

**DOI:** 10.1101/2023.09.18.23295755

**Authors:** Karina Maharjan, Rajan Paudel, Kshitij Kunwar

**Author notes:** Corresponding author: (KK).

## Abstract

People of all age groups are engaged with various screen-based devices. Use of screen-based media devices beyond duration considered problematic has become a problem worldwide. Yet, undergraduate students are engaged in various screen-based activities for academic works and entertainment which can influence their sleep.

A descriptive cross-sectional study design was used to collect information from 384 undergraduate students using a web-based questionnaire and convenience sampling technique. The data collected was analyzed using IBM SPSS Statistics 20.0. Descriptive and regression analyses were done to explore factors that influence screen-based media use.

The average duration of screen-based media use by the participants was found to be 7.12 hours per day. Factors such as sex of the participants, the use of screen-based media during major mealtimes and home-specific rules limiting screen-based media use were found to be associated with screen-based media use. Only 45.3% of participants got the recommended hours of sleep (7 or more hours). Around 9.4% of the participants had long sleep onset latency (more than 45 minutes). Participants who used screen-based media devices during night before sleep were more likely to get insufficient sleep (OR (Odds Ratio) 3.668, 95% CI (Confidence Interval) (1.784-7.539)).

A significant association was found between sex of the participants, use of screen-based media devices during major meal times and home rules limiting screen-based media use with the use of screen-based media for five hours or more per day. Screen-based media use and screen-based media use during night influence the quantity of sleep. Overuse of screen-based media is degrading the quality of sleep among undergraduate health students and is an alarming problem. There is a need to promote good sleeping habits through the reduction of this modifiable behaviour, which could be done by promoting home-based rules about screen-based media devices use, especially at night before sleep.

## Introduction

In the last decade there has been a sharp increase in availability and use of screen-based media devices such as television, laptop, smartphones, videogame consoles and computers and as a result people are exposed to unintentional artificial light [1]. Having access to these technologies has become an important part of everyday life. Most of the adults have at least one electronic media device in their bedroom. These electronic devices play an important part to support education of students and have also become a medium of entertainment. Nowadays people spend most of their leisure time using screen-based media devices. Not only during leisure time but they also tend to use it alongside performing any other work. College students are found engaged with various screen-based devices. They are engaged in various screen-based media related activities for the purpose of entertainment or works related to academics. Also, those devices are used to attend classes remotely, complete various assignments and assignments and reading materials are also distributed in forms that can be accessed through screen-based devices. Academic works are also increasing screen time of college students. It is seen that nine out of every ten college students use laptop or phone on a regular basis [2].

Use of screen-based media devices above recommended duration has become a problem worldwide. People of all age groups including children, adolescents, adults or elderly are engaged in various screen-based activities. While waking a teenager in the morning, there is higher odds that we will find a cell phone under their pillow. It was probably the last thing that they used before sleeping and probably the first thing that they will touch after waking up [3]. Screen-based media use is affected by various factors that may be individual factors i.e. gender, physical activity, home physical factors i.e. availability, location and accessibility of devices and social factors i.e. rules and limitation, TV co-viewing [4]. Screen time usage among students is due to academic requirement and need for being connected socially through electronic devices [2].

A study on technology use among college students found that students are found spending an average of 12 hours daily in some types of media among which nine and a half hours are spent with their gadgets like mobile devices, gaming devices, computers, etc [5]. Mostly people are awake 16 to 18 hours a day, among which spending 12 hours in electronic devices is a lot.

Sleep is an important part of human life. It is estimated that humans spend a third of their lifetime sleeping or attempting to do so [6]. Sufficient sleep duration is required for healthy growth and development of the mind and body especially for children and adolescents. College students should get about seven to nine hours of sleep [7]. Getting required amount of sleep helps improve grades, lowers the risk of obesity, improves memory, improves mood.

Active use of screen-based media among college going students especially night time use affects sleep in a negative way, it can be a factor for short sleep duration, disturbed sleep, increased sleep onset latency and delayed bed, wake up time. It causes sleep deprivation among them. College student may become sleep deprived due to various factors such as lighting, sleep environment, medications, body pain, aches, stress, shift based work [8]. Among these factors screen time and excessive use of light emitting device are common factors [9]. Surveys have shown the association of screen-based media use with sleep [6,9–12]. Sleep quality is found negatively correlated with hours spent on electronic devices [13]. Use of media before sleep is an ascribed factor for insufficient sleep among younger people and adult [14]. Studies have shown adverse association between screen-based media use and sleep health where delayed bedtime and shorter total sleep have been found to be most consistently related to media use [15–17].

Poor sleep can negatively affect academic performance. Inadequate sleep is also associated with cardiovascular, inflammatory and metabolic consequences. Insufficient sleep to an individual is shown to be associated with impaired cellular responses, depression, anxiety and obesity [6]. Therefore, this study aimed and has determined screen-based media device use, its associated factors and sleep outcomes among undergraduate health students from Kathmandu valley, Nepal.

## Methods

### Study design and setting including study site, design, sample size and sampling

The descriptive cross-sectional design was designed to assess the screen-based media device use, its associated factors and sleep outcomes among the undergraduate students of health background studying in colleges within Kathmandu valley (Kathmandu, Bhaktapur and Lalitpur districts). The respondents included Bachelor of Medicine and Bachelor of Surgery (MBBS), Bachelor of Dental Surgery (BDS), Bachelor in Public Health (BPH), Bachelor of Science in Medical Laboratory Technology (BSc. MLT), Bachelor in Pharmacy (B. Pharm), Bachelor of Science in Nursing (Bsc. Nursing), Bachelor of Nursing Science (BNS) and Bachelor in Optometry (B. optom), BPT (Bachelor in Physiotherapy), BASLP (Bachelor of Audiology & Speech Language Pathology) students. Data was collected from the undergraduates remotely by administering web-based questionnaire considering the risk of infection from COVID-19 by mobilizing the students’ network online.

The sample size was calculated using formula, n=Z^2p(1-p)/e^2 and sample size was calculated to be 347, assuming 10% non-response rate it was fixed as 382.

### Data collection

The questionnaire was adopted from previous studies Questionnaire regarding sleep was adapted based on Holly Scott [12] and questionnaire regarding screen-based media use and factors associated were adapted from Granich et.al [4]. The questionnaire had four sections: socio-demographic information, home media environment, screen time and sleep outcomes. The main outcome variable were screen time and factors affecting screen-based media use. Screen time referred to the total hours spent using screen-based media devices. Pretesting was carried out among 40 undergraduate students. The data was collected from 15-JAN-2021 to 28-FEB-2021.

Ethical approval for the study was obtained from the Institutional Review Board (IRB), Maharajgunj Medical Campus, Institute of Medicine, Tribhuwan University, Kathmandu, Nepal (Approval letter reference number: 201(6-11) E2 077/078). Participants were informed about the process, objective of the study, their privacy and confidentiality of the information they provide. Participants provided informed consent in the same web-based form before they could take part in the study by answering a question related to agreeing or disagreeing to take part in the study which they could voluntarily choose as “Yes” or “No”.

### Data processing and analysis

Data collected online was downloaded and Microsoft Excel was used to clean the data and was exported to IBM SPSS Statistics 20.0 for further coding and analysis. Information of independent variables were analysed using descriptive statistics namely frequency, percentage, median.

Inferential statistics using Chi-square test was done to find out the association between screen-based media use and independent variables and logistic regression was performed to determine the strength of association. Confidence interval of 95% was used and p-value <0.05 was considered statistically significant.

## Results

Table 1 shows the demographic characteristics 384 participants of the study which included 41.1% male and 58.9% female participants. The median age of the participants was 22 years.

The household media environment plays role in screen-based media behaviour. It was found that all participants come with a household of more than three media device of some kind and 79.4% participants had their own screen-based media devices as shown in Table 2. It was found that 21.6% of the participant’s household had some rules limiting the screen-based media use such as allocated screen time and in 39.6% participant’s homes, screen-based media devices were being viewed during major meals. Participant’s household with 3 or more variety of screen-based media device among the five types taken under this study was found to be 92.4%.

**Table 1:**
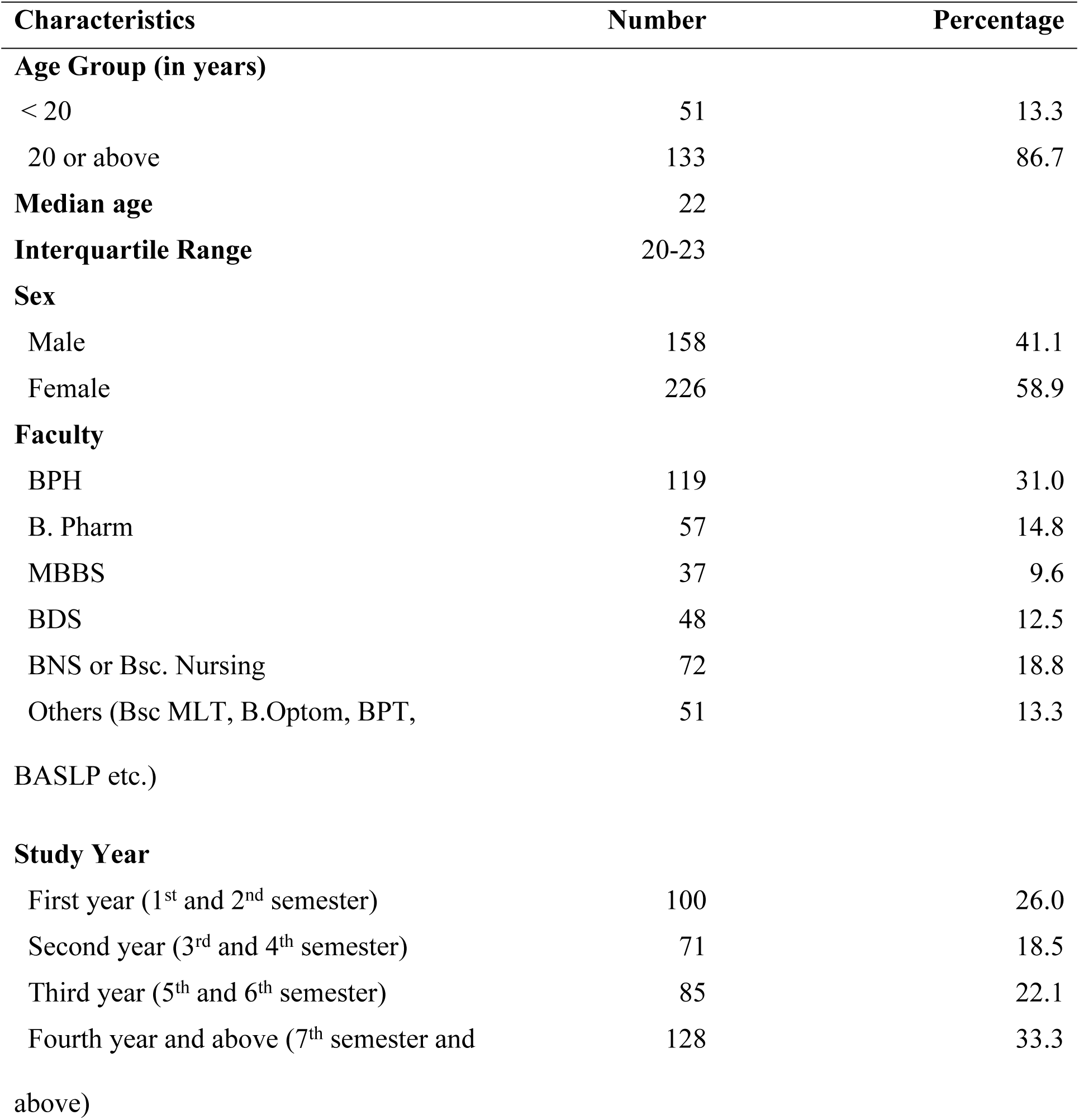
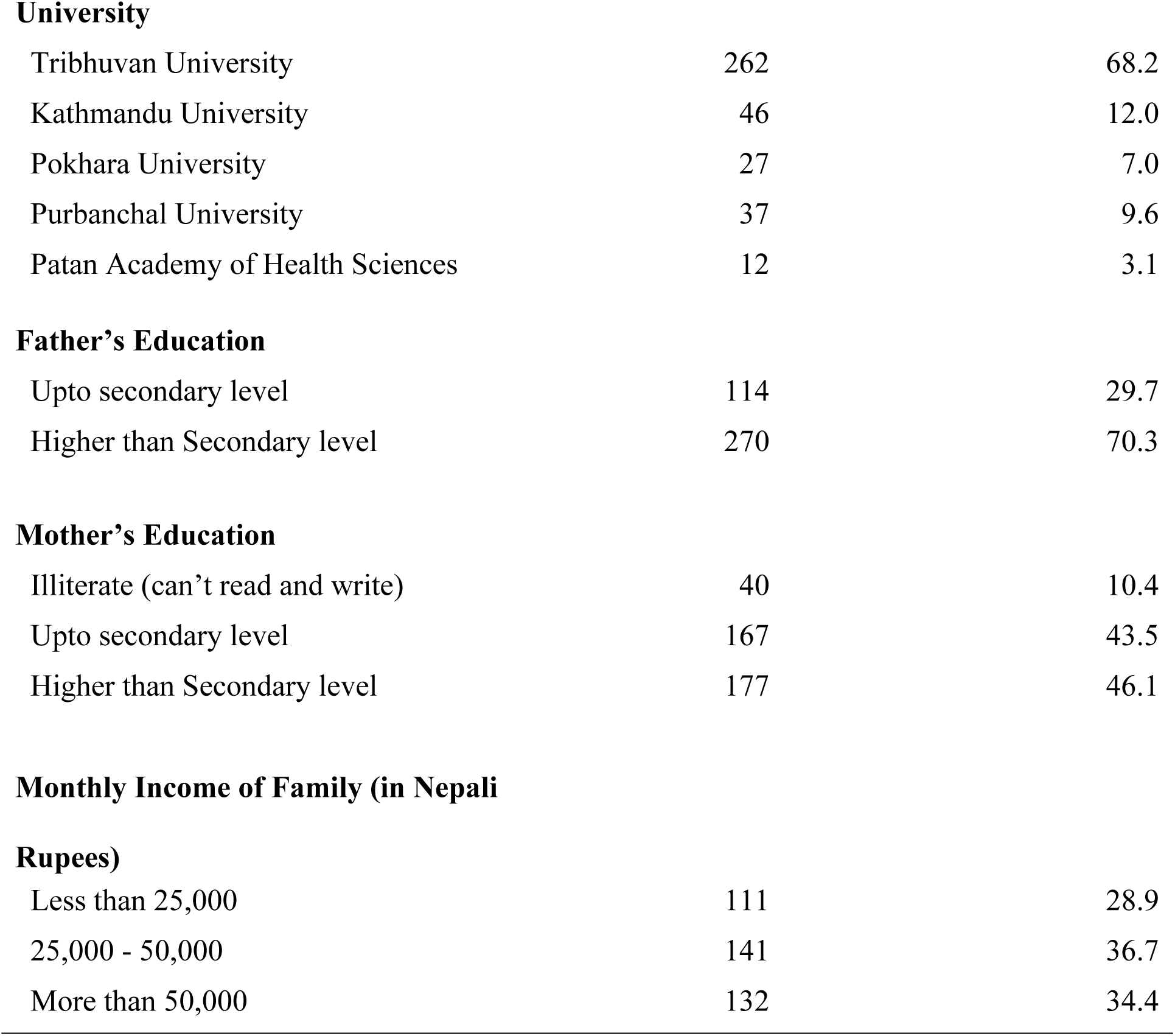
Demographic characteristics of participants (N=384)

| Characteristics | Number | Percentage |
| --- | --- | --- |
| **Age Group (in years)** |  |  |
| < 20 | 51 | 13.3 |
| 20 or above | 133 | 86.7 |
| **Median age** | 22 |  |
| **Interquartile Range** | 20-23 |  |
| **Sex** |  |  |
| Male | 158 | 41.1 |
| Female | 226 | 58.9 |
| **Faculty** |  |  |
| BPH | 119 | 31.0 |
| B. Pharm | 57 | 14.8 |
| MBBS | 37 | 9.6 |
| BDS | 48 | 12.5 |
| BNS or Bsc. Nursing | 72 | 18.8 |
| Others (Bsc MLT, B.Optom, BPT, BASLP etc.) | 51 | 13.3 |
| **Study Year** |  |  |
| First year (1<sup>st</sup> and 2<sup>nd</sup> semester) | 100 | 26.0 |
| Second year (3<sup>rd</sup> and 4<sup>th</sup> semester) | 71 | 18.5 |
| Third year (5<sup>th</sup> and 6<sup>th</sup> semester) | 85 | 22.1 |
| Fourth year and above (7<sup>th</sup> semester and above) | 128 | 33.3 |
| **University** |  |  |
| Tribhuvan University | 262 | 68.2 |
| Kathmandu University | 46 | 12.0 |
| Pokhara University | 27 | 7.0 |
| Purbanchal University | 37 | 9.6 |
| Patan Academy of Health Sciences | 12 | 3.1 |
| **Father's Education** |  |  |
| Upto secondary level | 114 | 29.7 |
| Higher than Secondary level | 270 | 70.3 |
| **Mother's Education** |  |  |
| Illiterate (can't read and write) | 40 | 10.4 |
| Upto secondary level | 167 | 43.5 |
| Higher than Secondary level | 177 | 46.1 |
| **Monthly Income of Family (in Nepali****Rupees)** |  |  |
| Less than 25,000 | 111 | 28.9 |
| 25,000 - 50,000 | 141 | 36.7 |
| More than 50,000 | 132 | 34.4 |

**Table 2:** Household media environment and screen-based media use (N=384)

| Household media environment and screen-based media use | Number | Percentage |
| --- | --- | --- |
| Have internet connection | 349 | 90.9 |
| Have access to TV channels | 320 | 83.3 |
| Have own screen-based media device | 305 | 79.4 |
| Presence of home Rules/Regulations about media use | 83 | 21.6 |
| Use screen-based media during major meals (breakfast, lunch, dinner) | 152 | 39.6 |
| Number of types of screen-based media devices present in the household |  |  |
| Less than 3 types | 29 | 7.6 |
| Equal to or more than 3 types | 355 | 92.4 |
| Number of types of screen-based media device owned by the respondent |  |  |
| None | 79 | 20.6 |
| One type | 90 | 23.4 |
| Two or more types | 215 | 56 |
| Average screen-based media use in 24 hours |  |  |
| ≥5 hours | 294 | 76.6 |
| <5 hours | 90 | 23.4 |
| Average duration of screen-based media use per 24 hours | 7.12 hours |  |
| Screen-based media use before bedtime |  |  |
| Doesn't use screen-based media | 43 | 11.2 |
| Uses screen-based media | 341 | 88.8 |
| **Duration of screen-based media use before bedtime** |  |  |
| ≤ 1 hour | 178 | 46.3 |
| 1-2 hour | 117 | 30.5 |
| >2 hour | 46 | 12.0 |

The major study variable of this research was screen-based media use among undergraduate students of the study area. Participants were asked total hours of use of screen-based media devices in 24 hours. The responses were then classified as <5 hours and ≥ 5 hours of use, on the basis of association seen with screen-based media use for more than or equals to five hours with computer vision syndrome.

Table 2 shows that 76.6 % participants spend more than or equals to 5 hours of their time being engaged in some kind of screen-based media activity in 24 hours. The average hours of screen-based media device use among the study participants was 7.12 hours per day (24 hours). Majority (69.5%) participants responded that they prefer to use smartphones over other screen-based medias. As for the purpose of use, majority (46.1%) of the participants reported using screen-based media devices for entertaining themselves.

Table 3 shows the sleep related outcomes of the participants with average sleep of 6.72 hours. Sleep onset latency refers to the time required for an individual to fall asleep, it was found that 9.4% participants have long SOL (Sleep Onset Latency) (more than 45 minutes).

**Table 3:** Sleep outcomes of participants (N=384)

| Sleep Outcomes | Number | Percentage |
| --- | --- | --- |
| **Sleep time** |  |  |
| Before 11 PM | 214 | 55.7 |
| 11 PM and after | 170 | 44.3 |
| **Sleep Onset Latency (SOL)** |  |  |
| ≤ 45 minutes | 348 | 90.6 |
| > 45 minutes | 36 | 9.4 |
| **Total sleep duration** |  |  |
| <7 hours | 210 | 54.7 |
| ≥ 7 hours | 174 | 45.3 |
| Average hours of sleep | 6.72 hours |  |
| Range of total hours of sleep | 2-11 hours |  |

Results from the study showed that screen-based media use was associated with sex of the participants, mealtime viewing of the screen-based media and rules limiting screen-based media use as shown in Table 4.

**Table 4:** Association between variables and screen-based media use.

| Characteristics | Screen-based media use |  | p-value |
| --- | --- | --- | --- |
|  | ≥ 5 hours (%) | < 5 hours (%) |  |
| **Age in years** |  |  |  |
| <20 | 38 (74.5) | 13 (25.5) | 0.710 |
| 20 and above | 256 (76.9) | 77 (23.1) |  |
| **Sex** |  |  |  |
| Male | 138 (87.3) | 20 (12.7) | <0.001 |
| Female | 156 (69) | 70 (31) |  |
| **Own personal device** |  |  |  |
| Yes | 237 (77.7) | 68 (22.3) | 0.300 |
| No | 57 (72.2) | 22 (27.8) |  |
| **Has internet Access** |  |  |  |
| Yes | 264 (75.6) | 85 (24.4) | 0.187 |
| No | 30 (85.7) | 5 (14.3) |  |
| **Availability of TV channels** |  |  |  |
| Available | 240 (75) | 80 (25) | 0.110 |
| Not available | 54 (84.4) | 10 (15.6) |  |
| **Uses screen-based media device during major meal time** |  |  |  |
| Yes | 126 (82.9) | 26 (17.1) | **0.019** |
| No | 168 (72.4) | 64 (27.6) |  |
| **Presence of Rule about using screen-based media device at household** |  |  |  |
| Yes | 56 (67.5) | 27 (17.1) | **0.029** |
| No | 238 (79.1) | 63 (20.9) |  |
| **Uses screen-based media device in bedroom** |  |  |  |
| Yes | 156 (77.2) | 46 (22.8) | 0.746 |
| No | 138(75.8) | 44 (24.2) |  |
| **Number of types of devices owned by the respondent** |  |  |  |
| Less than three types | 25 (86.2) | 4 (13.8) | 0.210 |
| Three or more types | 269 (75.8) | 86 (24.2) |  |

This study found that there was no association of personal media ownership and electronic media device in bedroom with night time screen-based media use.

The results of logistic regression analyses are shown in Table 5. Female participants were less likely to have more than or equal to 5 hours screen time (AOR (Adjusted Odds Ratio) 0.356, 95% CI (1.200-0.635)) than male participants. Participant who do not view screen-based media devices while having meal was less likely to have screen time of more than or equal to 5 hours (AOR 0.474, 95% CI (0.277 – 0.812)) then those who don’t. Likewise, participants not having home rules limiting screen time were more likely to have screen time of more than or equal to 5 hours (AOR 1.788, 95% CI (1.013-3.155)) than those who have rules.

**Table 5:**
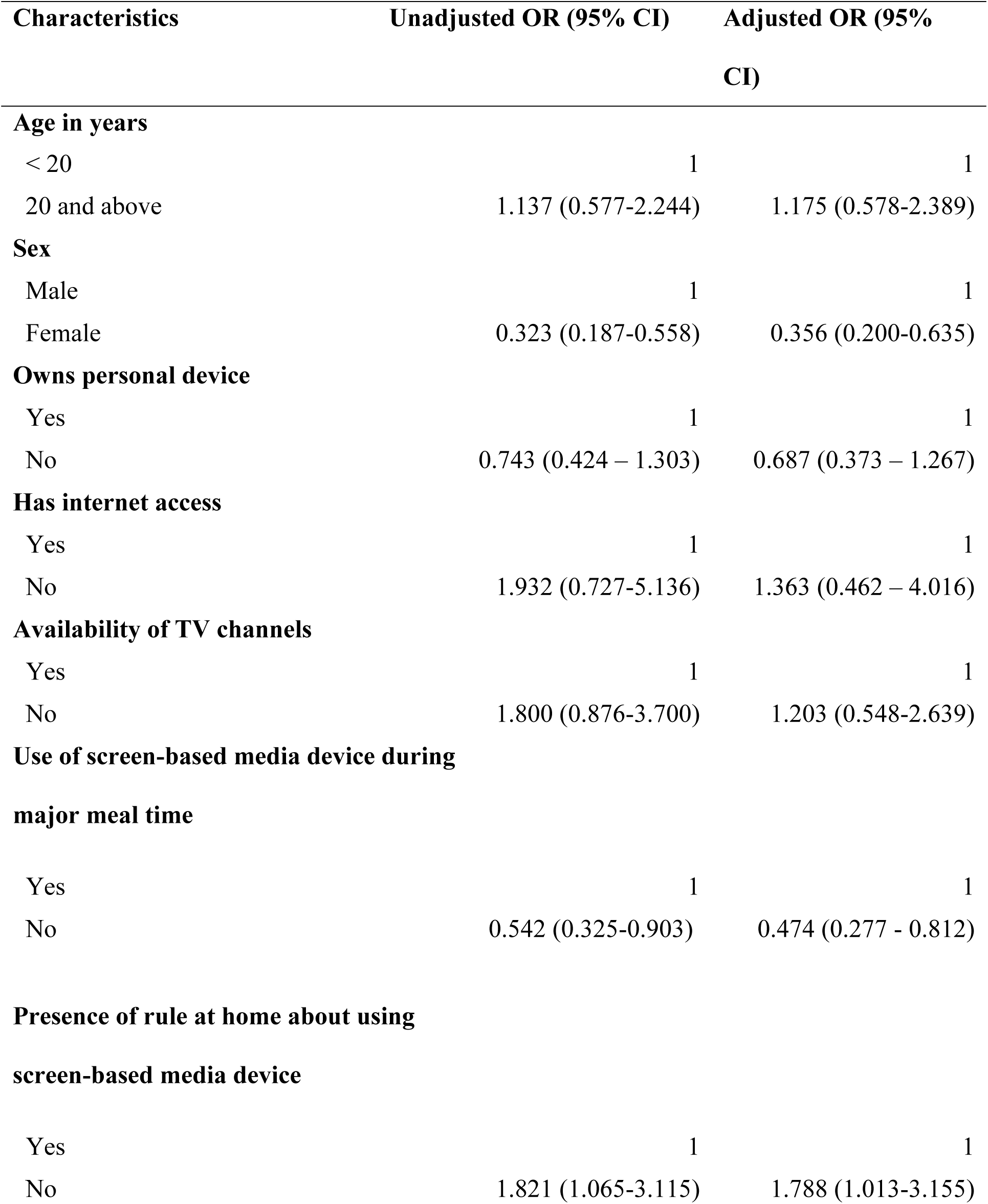
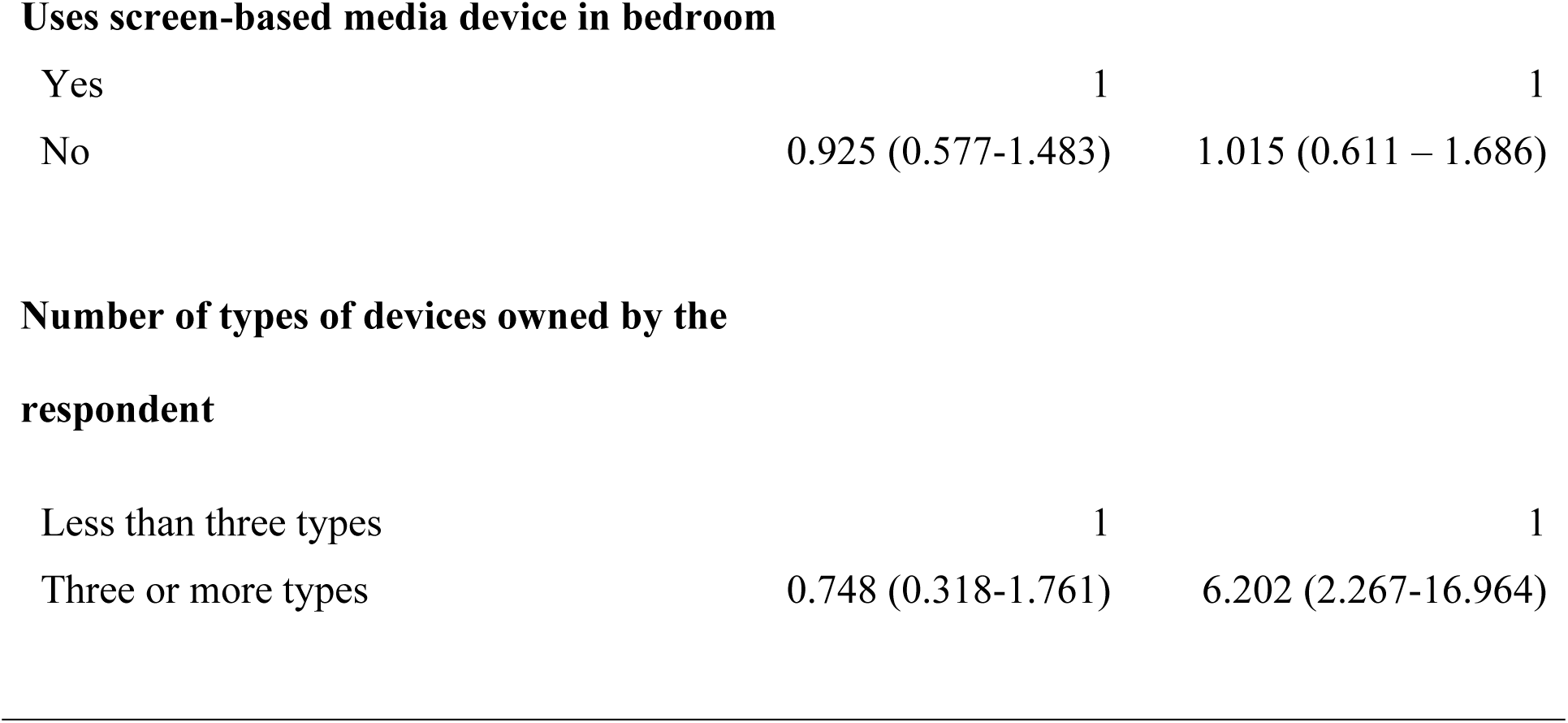
Logistic regression for factors of screen use for more than recommended hours (≥ 5 hours)

| Characteristics | Unadjusted OR (95% CI) | Adjusted OR (95% CI) |
| --- | --- | --- |
| **Age in years** |  |  |
| < 20 | 1 | 1 |
| 20 and above | 1.137 (0.577-2.244) | 1.175 (0.578-2.389) |
| **Sex** |  |  |
| Male | 1 | 1 |
| Female | 0.323 (0.187-0.558) | 0.356 (0.200-0.635) |
| **Owns personal device** |  |  |
| Yes | 1 | 1 |
| No | 0.743 (0.424 – 1.303) | 0.687 (0.373 – 1.267) |
| **Has internet access** |  |  |
| Yes | 1 | 1 |
| No | 1.932 (0.727-5.136) | 1.363 (0.462 – 4.016) |
| **Availability of TV channels** |  |  |
| Yes | 1 | 1 |
| No | 1.800 (0.876-3.700) | 1.203 (0.548-2.639) |
| **Use of screen-based media device during major meal time** |  |  |
| Yes | 1 | 1 |
| No | 0.542 (0.325-0.903) | 0.474 (0.277 - 0.812) |
| **Presence of rule at home about using screen-based media device** |  |  |
| Yes | 1 | 1 |
| No | 1.821 (1.065-3.115) | 1.788 (1.013-3.155) |
| **Uses screen-based media device in bedroom** |  |  |
| Yes | 1 | 1 |
| No | 0.925 (0.577-1.483) | 1.015 (0.611 – 1.686) |
| **Number of types of devices owned by the respondent** |  |  |
| Less than three types | 1 | 1 |
| Three or more types | 0.748 (0.318-1.761) | 6.202 (2.267-16.964) |

Regression analysis presented in Table 6 showed that participants who use screen-based media devices for **≥** 5 hours a day were more likely to get inadequate sleep (AOR 3.898, 95% CI (2.325-6.534)) than those who use screen-based media for less than 5 hours a day. Likewise, participants who view screen-based media device before sleeping were more likely to get inadequate sleep (AOR 3.668, 95% CI (1.784-7.539)) than those who do not view screen-based media device before sleep.

**Table 6:** Logistic regression for factors of inadequate sleep (<7 hours)

| Characteristics | Unadjusted OR (95% CI) | Adjusted OR (95% CI) |
| --- | --- | --- |
| **Duration of use of screen-based media** |  |  |
| $\geq 5$ hours | 3.847 (2.313 – 6.398) | 3.898 (2.325-6.534) |
| < 5 hours | 1 | 1 |
| **Night time screen-based media use** |  |  |
| Yes | 3.567 (1.776-7.205) | 3.668 (1.784-7.539) |
| No | 1 | 1 |

## Discussion

This study was carried out to find out the screen-based media use, factors associated with it and sleep related outcomes among health undergraduate students studying in colleges of Kathmandu valley. The study has assessed the present scenario of screen-based media use among undergraduate students and their sleep outcomes.

This study found that undergraduate students spend an average of 7.2 hours in a day using some form of screen-based media device which is similar to findings from a study done by Kaiser Family Foundation that showed average use screen-based media device for 7.5 hours per day among young adolescents [3]. Similar study done by Yen-Sen Lee et al, found that the students spend an average of 8.3 hours in a day using screen-based devices [13]. This study showed that most of the participants are engaged in screen-based activity during evening time. A study by Angela Lynn Sargent on screen time and sleep condition among selected college students showed that 75% students reported always use of electronic media device one hour before bedtime [18]. Similarly, this study showed that 88.8% participants reported using screen-based media device before sleeping.

A study by Granich and Kaiser Family Foundation studied the relationship of household screen-based media and screen-based media use among adolescents. Both study have concluded that there is positive association between availability and accessibility of media devices with time spent on screen-based media use [3,4]. In contrast to this study, the current study showed no association between accessibility and availability of screen-based media device with its use. Though, significant association was seen between having home rules limiting screen-based media use and meal time viewing of screen-based media devices with screen-based media use among the study participants. The percentage of individuals with an internet connection was recorded to be 36.7% in 2021 [19]. In contrast to this, the household with internet access were measured to be 90.9% in this study. This could be due to the fact that the sample was based on the urban population of Kathmandu valley. The entire participant’s household owned more than three screen-based media devices of any kind and 79.4% participants had their own screen-based media device, the most commonly owned device was smartphones.

A study by Angela Lynn Sargent on screen time and sleep condition among selected college students showed that students on average get 6.9 hours of sleep. It also showed that 65.5% of the participants reported getting recommended 7 hours or more sleep. The study found no statistically significant difference regarding screen use before bedtime and participants hours of sleep [18]. A study carried out to see the association between portable screen-based media use and sleep outcome showed that participants who had access to media device at night were more likely to have inadequate sleep quantity, OR 1.79, 95% CI 1.39-2.31 [11]. Similarly, the current study found out that the participants got an average of 6.72 hours of sleep and also found that only 45.3% participants got recommended hours of sleep. Similar to the previous study, significant association was found between night time screen-based media use and participants sleep. A study by Nowell SB et al studied the relationship between social media use and sleep quality of undergraduate nursing students showed that sleep duration among students ranges from 4 to 10 hours, the study also showed that electronic media use correlates with poor sleep quality [20]. This study shows that the sleep duration of participants ranges from 2 to 11 hours.

The data presented in the study were self-reported and partly depend on the participant recall ability thus they may subject to recall bias. This study is limited to factors affecting screen-based media use among undergraduates, sleep outcomes and it does not address any health effects due to sleep inadequacy. There are many physical, psychological, behavioural and emotional aspects of screen time as well but the variables that were studied in this study were focused only on sleep related outcomes. Also, there are many other factors affecting sleep outcomes of an individual beside screen-based media use but only screen-based media was studied as a factor affecting sleep.

## Conclusion

This study showed that the participants on average use screen-based media devices for 7.2 hours per day and only 45.3 % participants got recommended hours of sleep (7 or more hours). Around 9.4% of the participants had long sleep onset latency (more than 45 minutes). Participants who use screen-based media devices during night time before sleep were more likely to get insufficient sleep (AOR 3.668, 95% CI (1.784-7.539)).

Significant association was found between screen-based media use and variables such as sex of participant, use of screen-based media device during major meal time and home rules limiting screen-based media. Significant association was found between sleep adequacy with screen-based media use and night time screen-based media use. Thus, this research suggests the need to limit the use of screen-based media devices to promote quality of sleep. Overuse of screen-based media stands as an alarming problem among undergraduates and this can have direct influence on their physical and mental health. Limiting the use of screen-based media devices through the promotion of home-specific rules about screen-based media use especially about the use at night time before sleep can help undergraduate students get the recommended amount of sleep.

## Data Availability

All data underlying the study findings are fully available without restriction.

## Acknowledgement

We would like to thank all respondents for their participation and academics of Institute of medicine for their academic guidance and support for this study.

## Author’s contribution

Conceptualization: KM,

Data curation: KM, KK

Formal analysis: KM,

Investigation: KM,

Methodology: KM, RP,

Project administration: KM,

Resources: KM,

Supervision: RP,

Validation: RP, KK,

Writing – original draft: KM,

Writing – reviewing & editing: KK, KM, RP

## Supporting information

**S1 Dataset. Study dataset and data labels**

**S2 Appendix. Study questionnaire**

**S3 Appendix. Results of regression analysis**

**S4 File. Institutional Review Committee approval letter**

